# Work-related road traffic crashes: emergence of new modes of personal journey. Analysis based on data from a register of road traffic crashes

**DOI:** 10.1101/2024.04.04.24305326

**Authors:** Emmanuel Fort, Nicolas Connesson, Julien Brière, Amina Ndiaye, Blandine Gadegbeku, Barbara Charbotel

## Abstract

**Introduction:** According to the 2018–2019 People Mobility Survey, work-related journeys (commuting and on-duty journeys) account for approximately 25% of all journeys. The use of non-motorized (nm) and motorized (m) personal mobility devices (PMDs) has steadily increased since their introduction into the French market in the last decade.

**Objective:** This study aimed to describe the characteristics of work-related road crashes and their evolution since the introduction of new PMDs in France and the increase in the use of scooters.

**Materials and methods:** This was a retrospective, cross-sectional study using data from the Rhône Road Trauma Registry. Data were collected from 2015 to 2020. We included the data for the victims aged 18–70 years who were injured in work-related road crashes.

**Results:** We identified 11,296 individuals aged 18–70 years who experienced work-related road crashes. An injury report was provided for a total of 11,277 patients. A total of 546 passengers and 78 drivers of other motorized vehicles (buses/trams, construction equipment, and tractors) were excluded from the analysis. Seven patients died at the time of the crash, and seven died after hospitalization. Of the 10,653 (94.4%) victims, there were pedestrians (5.1%), or riders of bicycles (16.9%), scooters (3.8%), other PMDs (roller blades, skateboards, monowheels, gyropods, and hoverboards; 0.4%) and motorized two-wheeler (21.4%), or drivers of car (45.3%), and truck (1.5%). More than half of the scooter riders and 80% of other PMD riders were men. More than 60% of other PMD riders and 53% of scooter riders were under 34 years of age. Most scooter road crashes occurred during commuting (95.6%). Sixty-five percent of the scooter crashes and 50% of other PMD crashes did not have opponents. Overall, one-quarter of the victims experienced crashes without opponents. Most scooter riders had injuries to their upper limbs (59.2%), lower limbs (46.8%), face (21.2%) or head (17.9%).

**Discussion:** Most work-related road crashes were of low or moderate severity (97.5%; Maximum Abbreviated Injury Scale, MAIS score < 3). The frequency and severity of injuries among scooter and other PMD users were similar. Scooter opponents were rarely observed in pedestrian injuries (12/575). Most scooter- and other PMD-crashes did not have opponents.

**Conclusion:** Many head injuries could be prevented with more widespread use of helmets, among scooter- and other PMD users and bicycle users.

**What is already known on this topic:**

- The use of personal mobility devices (PMDs) has steadily increased in France.
- Work-related journeys (commuting and on-duty journeys) account for approximately 25% of all journeys

**What this study adds:**

- We describe work-related road crashes’ characteristics since PMDs’ introduction.
- Work-related road crashes involving scooters or other PMDs riders are of low severity.
- Most scooter riders had injuries to their upper limbs (59.2%), lower limbs (46.8%), face (21.2%) or head (17.9%).

**How this study might affect research, practice or policy:**

- Many head injuries could be prevented with more widespread helmet use.
- Companies can take preventive actions to ensure that employees are better informed.

## 1. Introduction

While a large proportion of travel in France involves private journeys (leisure or other), work-related journeys (home-work and on-duty) account for approximately 25% according to the 2018–2019 People Mobility Survey [1]. The main modes of transportation for commuting journeys were cars (72%), public transport (12%), walking (9%), cycling (3%), and motorized two-wheelers (2%); other modes accounted for less than 1%. According to the National Transport and Mobility Survey, work-related journeys accounted for approximately 27% of all journeys in 2008 [2]. The 2016 National Survey of Environmental Behavior [3] showed that 25% of journeys were mainly for commuting. According to the Sumer surveys in France, 25% of employees drive on public roads as part of their work [4,5]. This proportion has remained constant since the surveys were conducted (2003, 2010, and 2017).

The mode of transport to reach work depends on several factors [3]. The distance to work, availability of transportation, car ownership, household size, parking, and cost are determining factors in choosing the travel mode(s). In addition, there is a significant difference between the types of territory; walking, cycling, and other soft modes of transport are preferred in Paris and its suburbs, as well as in large cities and urban areas (those with 100,000–10,000,000 inhabitants).

A personal mobility device (PMD) is a class of compact micromobility vehicle for transporting an individual. It can be separated in 2 subgroups: the non-motorized personal mobility devices (nmPMDs) which can be typically scooter, roller, skateboard; and motorized personal mobility devices (mPMDs) which include electric scooter, skateboards, kick scooters, self-balancing unicycles and Segways, as well as gasoline-fueled motorized scooters or skateboards, typically using two-stroke engines of less than 49 cc (3.0 cu in) displacement and which can’t ride at speeds that do normally exceed 25 km/h (16 mph) or sometimes 20km/h in some countries. The use of PMD has been steadily increasing, mostly since the introduction of electric scooter into the market in France in the last decade, as well as in other cities globally [6]. These modes of transport can be referred to as micromobility that combines all easy modes of transport that allow their users to make hybrid use of the vehicle either as a pedestrian or as a vehicle occupant [7]. It represents between 8–15% of daily journeys of less than 8 km, which today account for 50–60% of the distance traveled in China, the European Union, and the USA, and replaces 20% of public transport journeys (in addition to bridging the gap between the first and last kilometers) as well as bicycle, moped, scooter, or on-foot journeys [8].

The introduction of electric scooter sharing (in June 2018 in France) has accelerated the use of electric scooters for journeys. It has been observed in all industrialized countries, mainly in large metropolises [9]. In overall, the development of PMDs is growing, primarily in urban areas, for several reasons. First, their use could replace motorized vehicles, such as cars or motorized two-wheelers. Second, they reduce air pollution; therefore, their use is favored in cities with reserved lanes (for bicycles and PMDs) or shared lanes (usually for bicycles, PMDs, bus and taxi). Finally, the increase in the cost of fossil energy favors the development of PMDs. In fact, it appears that most walking and public transport journeys are replaced by trips using PMDs [10]. Electric scooters replace walking or biking for the last mile of a journey [11], potentially reducing the overall physical activity of the population [12]. The introduction of large low-emission zones (ZFE), where vehicle speeds are limited to 30 km/h, is a new incentive to use PMDs. The COVID-19 pandemic has increased the use of PMDs, particularly by encouraging social distancing [13]. Thus, public transport users have abandoned subways, tramways, or buses, where distances between people can be too small, for the use of PMDs. However, some authors have highlighted problems caused by electric scooters, both regarding the safety of other vulnerable users and the impact on the environment during the production of scooters, especially batteries, which cause pollution [14].

In Vienna, Austria, Laa and Leth [15] showed that users were shifting to electric scooters, followed by buses and street cars, to replace walking. Electric scooters are used in combination with other modes of transport. A study from the USA on users of the shared-bike service in Washington, D.C. found that 35% of occasional users reported using it as a substitute for public transport [16]. In Belgium, nearly 50% of customers reported replacing one or more modes of transport with electric scooters [17].

PMDs now represent a significant part of urban and peri-urban transport modes. In France, 22% of respondents have used them at least once, and 11% are regular users (at least once a month) [18]. The electric scooters are the most-used mPMD, followed to a lesser extent by the gyro-mot, electric skateboard, gyropod, and hoverboard. One of two daily users of mPMD uses a self-service electric scooter. In Greece, 40% of the users mentioned using electric scooters for work purposes [9]. These electric vehicles offer new opportunities to users in terms of travel speed and less effort than their non-electric counterparts; however, they cause problems with public space sharing and road safety [18].

Many regulations have been implemented and experiments have been conducted in cities and countries to better control the use of electric scooters [6] and reduce the injuries associated with them, such as restrictions on hiring and using electric scooters at night [19], wearing helmets, adhering to speed limits, and traffic zones [6].

Many studies have found an increase in injuries associated with the use of electric-sharing scooters In USA, the estimated incidence of electric scooter injuries treated in emergency departments in the US nearly doubled between 2018 and 2019 [20]. In the United Kingdom, a case series demonstrated an increasing frequency of significant orthopedic injury associated with electric scooter use treated at a Level 1 Major Trauma Center over the course of two years [21] while a study in a trauma unit found a significant rise in electric scooter-related injuries seen between 2019 and 2020 [22]. A study about the patients attended on in an emergency room in Spain concluded that the popularization of electric scooters among the employed population has caused a high increased number of crashes in this range of age [22]. Finally, in Finland, [23] and in France [24] other studies have found an increase in injuries associated with the introduction and the use of electric-sharing scooters [25–29].

Thus, although this increase has not yet been quantified in mobility surveys, there has been an increase in road crashes for users. In a recent publication, the Rhône Registry reported 1,186 scooter crashes, resulting in 1,197 injured users and a 7.3-fold increase in the number of scooter crashes between 2018 and 2019 [30].

Therefore, this study aimed to describe the characteristics of victims injured during a work-related road crash and their evolution since the emergence of PMDs, more specifically the scooters. A secondary objective was to characterize and compare crashes involving scooters and other PMDs with the usual user categories.

## 2. Materials and Methods

### 2.1. Design

This retrospective cross-sectional study used data from the Rhône Road Trauma Registry.

### 2.2. The Rhône Road Trauma Registry

The Rhône Road Trauma Registry has been recording data prospectively since its inception in 1995 and covers the Rhône region (1.85 million inhabitants), including one of France’s largest cities (Lyon, 0.5 million inhabitants, 11,000 inhabitants/km^2^) within a metropolis (Lyon Metropolis, 1.4 million inhabitants). Individuals were included in the registry if they faced a road crash involving one or more road users (motorized or not) in the Rhône region and consulted one of the 245 public or private healthcare facilities (including 42 emergency departments and 20 intensive care units within levels I, II, and III trauma centers), including pre-hospital primary care services and forensic medicine institutes.

The Registry collects the data on demographics of each victim, their sustained injuries, and crashes. Patient information was prospectively collected from the time of the crash to hospital discharge from prehospital emergency care, emergency departments, intensive care units, and surgical units.

### 2.3. Study population

Data were collected from 2015 to 2020. The victims aged 18–70 years (legal age limit for working in France for workers) who were injured on the way to/from work or while working were included in the Rhône Registry database (Figure 1). Persons without an injury were excluded because they were out of the scope of our study, as were passengers in a vehicle (n=546) because they were not driver and drivers of buses/coaches/tramway (n=61) or other motorized vehicles (tractors, construction equipment, quad bikes; n=17) because they were few and with specific particularities that don’t allow relevant comparisons with other users (defined paths for buses and coaches).

**Figure 1:**
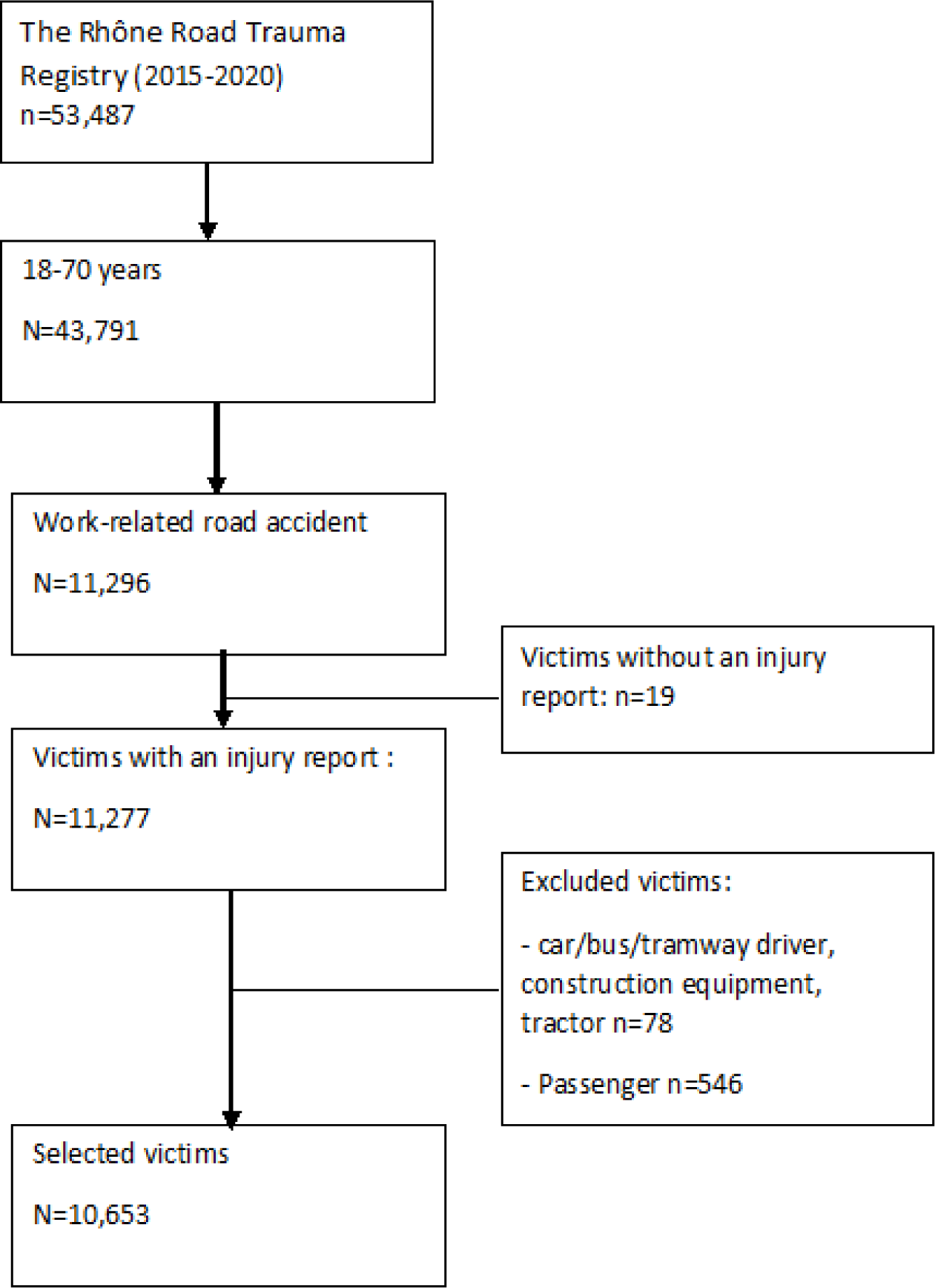
Flow chart.

### 2.4. Variables

The following variables were analyzed: sex, age, category of work journey (commuting or mission), category of road user, type of road network, opponent, time of crash, month of crash, day of crash, use of adequate safety equipment, hospitalization, injuries and death at the scene or post-crash.

### 2.5. Gravity of the injuries

Each injury was coded according to the Abbreviated Injury Scale (AIS), the AIS 2005 Revision, which comprises 2,000 codes divided into nine body areas [31,32]. Each code has an associated AIS severity score ranging from 1 (minor injury) to 6 (beyond treatment/death). As a patient may have multiple injuries in the same body part, the maximum AIS (MAIS) reports the greatest severity in each injury area. Similarly, the overall MAIS score is calculated to determine the victims’ overall severity of injuries. Finally, the New Injury Severity Score (NISS) was calculated from the three most severe injuries and was defined as the sum of the squares of the respective AIS [33,34]. It was then classified into 3 categories: minor trauma <9, moderate trauma (9–15), and severe trauma >=16.

### 2.6. Statistical analysis

A descriptive analysis was conducted for the overall data and by user category (for the main categories). Seven categories of users were identified: pedestrians, bicycle riders, scooter riders, riders of other PMDs (roller blades, skateboards, hoverboards, and monowheels), car/light commercial vehicle drivers, truck drivers, and motorized two-wheeler riders. We also considered these 7 categories into 2 subgroups: “soft mode transport users” (pedestrians, bicyclists, scooters, and other PMDs) and “motor vehicle users not authorized to use bicycle paths and lanes” (cars/light commercial vehicles, trucks, and motorized two-wheelers).

Firstly, we described the victims of the 2015-2020 register, grouped into 7 user categories according to socio-demographic characteristics (gender, age) and crash characteristics (type of commute, antagonist, month of crash,…), with comparisons between the 7 user categories as a whole and between users belonging to 2 sub-groups, then we describe and compare the same groups according to injury characteristics (MAIS, area of injury,…), and finally we studied the trends over the 2015-2019 period.

Categorical variables were compared by user category using the chi-square test, and quantitative variables, using analysis of variance (ANOVA). All analyses were performed using SAS software version 9.4.

## 3. Results

### 3.1. Description of the victims of work-related road crash

Between 2015 and 2020, 53,487 victims were recorded in the Rhône registry, of whom approximately 82% (n=43,791) were between 18 and 70 years of age. Approximately 26% (n=11,296) of crashes were related to commutes or duty journeys.

Of the 11,296 victims aged 18–70 years involved in work-related road traffic crashes, 11,277 had an injury report. Seven people died at the time of the crash and seven died after hospitalization.

We excluded 546 passengers and 78 drivers of other motorized vehicles (buses, tramways, construction equipment, and tractors) from the analysis according to the main categories of interest, resulting in 10,653 participants (94.4%). These were scooter riders (3.8%), other PMD riders (0.4%), bicycles riders (16.9%), motorized two-wheelers riders (21.4%), pedestrians (5.1%), and cars or light commercial vehicles (45.3%) and trucks (1.5%) drivers.

Nearly half of the scooter riders (54.8%) and more than three-quarters of other PMD riders were male (Table 1). Overall, most victims were male (60.9%), except for pedestrians (45.7%) and car occupants (47.1%).

**Table 1:** Socio-demographic and characteristics of the crash of victims injured in work-related crash, by user category (source the Rhône Road uma Registry, 2015-2020)

|  | Pedestrian<br>(n=575) | Bicycle rider<br>(n=1 908) | Scooter<br>rider<br>(n=429) | Other PMD<br>rider<br>(n=50) |  |  | Car driver<br>(n=5 111) | Truck driver<br>(n=166) | Motorized<br>two-wheelers<br>rider<br>(n=2 414) |  |  | All (n=10<br>653) |  |
| --- | --- | --- | --- | --- | --- | --- | --- | --- | --- | --- | --- | --- | --- |
|  | n (%) | n (%) | n (%) | n (%) | p-value<sup>1</sup> |  | n (%) | n (%) | n (%) | p-value<sup>2</sup> |  |  | p-value<sup>3</sup><br>(All) |
| **Sex** |  |  |  |  | <0.0001 |  |  |  |  | <0.0001 |  |  | <0.0001 |
| Women | 312 (54.3) | 664 (34.8) | 194 (45.2) | 10 (20.0) |  |  | 2703 (52.9) | 3 (1.8) | 277 (11.5) |  |  | 4163 (39.1) |  |
| Men | 263 (45.7) | 1244 (65.2) | 235 (54.8) | 40 (80.0) |  |  | 2408 (47.1) | 163 (98.2) | 2137 (88.5) |  |  | 6490 (60.9) |  |
| **Age categories** |  |  |  |  | <0.0001 |  |  |  |  | <0.0001 |  |  | <0.0001 |
| 18-24 years | 139 (24.2) | 355 (18.6) | 94 (21.9) | 15 (30.0) |  |  | 1208 (23.6) | 27 (16.3) | 678 (28.1) |  |  | 2516 (23.6) |  |
| 25-34 years | 140 (24.4) | 636 (33.3) | 132 (30.8) | 15 (30.0) |  |  | 1597 (31.2) | 42 (25.3) | 683 (28.3) |  |  | 3245 (30.5) |  |
| 35-44 years | 113 (19.6) | 426 (22.3) | 108 (25.2) | 12 (24.0) |  |  | 1149 (22.5) | 39 (23.5) | 499 (20.7) |  |  | 2346 (22.0) |  |
| 45-54 years | 108 (18.8) | 331 (17.4) | 77 (17.9) | 4 (8.0) |  |  | 832 (16.3) | 34 (20.5) | 388 (16.1) |  |  | 1774 (16.7) |  |
| 55-64 years | 69 (12.0) | 157 (8.2) | 17 (4.0) | 4 (8.0) |  |  | 317 (6.2) | 21 (12.6) | 164 (6.8) |  |  | 749 (7.0) |  |
| 65-70 years | 6 (1.0) | 3 (0.2) | 1 (0.2) | 0 (0) |  |  | 8 (0.2) | 3 (1.8) | 2 (0.1) |  |  | 23 (0.2) |  |
| **Type of work-related journey** |  |  |  |  | <0.0001 |  |  |  |  | <0.0001 |  |  | <0.0001 |
| Commuting | 456 (79.3) | 1742 (91.3) | 410 (95.6) | 43 (86.0) |  |  | 4565 (89.3) | 35 (21.1) | 2041 (84.5) |  |  | 9292 (87.2) |  |
| On duty | 119 (20.7) | 166 (8.7) | 19 (4.4) | 7 (14.0) |  |  | 546 (10.7) | 131 (78.9) | 373 (15.5) |  |  | 1361 (12.8) |  |
| **Opponent** |  |  |  |  | NA |  |  |  |  | NA |  |  | NA |
| None | 0 (0) | 909 (47.6) | 281 (65.5) | 26 (52.0) |  |  | 548 (10.7) | 48 (28.9) | 1015 (42.1) |  |  | 2827 (26.5) |  |
| Pedestrian | 0 (0) | 29 (1.5) | 3 (0.7) | 1 (2.0) |  |  | 7 (0.1) | 0 (0) | 11 (0.5) |  |  | 51 (0.5) |  |
| Other or unknown | 14 (2.4) | 19 (1.0) | 4 (0.9) | 0 (0) |  |  | 15 (0.3) | 0 (0) | 11 (0.5) |  |  | 63 (0.6) |  |
|  | n (%) | n (%) | n (%) | n (%) | p-value<sup>1</sup> |  | n (%) | n (%) | n (%) | p-value<sup>2</sup> |  |  | p-value<sup>3</sup><br>(All) |
| Car | 391 (68.0) | 646 (33.9) | 87 (20.3) | 16 (32.0) |  |  | 3414 (66.8) | 34 (20.5) | 1139 (47.2) |  |  | 5727 (53.8) |  |
| Fixed obstacle | 0 (0) | 164 (8.6) | 31 (7.2) | 3 (6.0) |  |  | 584 (11.4) | 43 (25.9) | 112 (4.6) |  |  | 937 (8.8) |  |
| Light commercial vehicle, truck,<br>bus, train/tram, tractor | 66 (11.5) | 63 (3.3) | 10 (2.3) | 1 (2.0) |  |  | 517 (10.1) | 40 (24.1) | 85 (3.5) |  |  | 782 (7.3) |  |
| Motorized two-wheelers | 52 (9.0) | 21 (1.1) | 5 (1.2) | 1 (2.0) |  |  | 23 (0.5) | 1 (0.6) | 31 (1.3) |  |  | 134 (1.3) |  |
| Bicycle | 52 (9.0) | 56 (2.9) | 8 (1.9) | 2 (4.0) |  |  | 3 (0.1) | 0 (0) | 10 (0.4) |  |  | 131 (1.2) |  |
| Rollerblade, skateboard | 0 (0) | 1 (0.05) | 0 (0) | 0 (0) |  |  | 0 (0) | 0 (0) | 0 (0) |  |  | 1 (0) |  |
| **Lighting** |  |  |  |  | 0.0009 |  |  |  |  | 0.5 |  |  | 0.002 |
| Day | 320 (65.4) | 1067 (74.1) | 218 (76.5) | 31 (75.6) |  |  | 3072 (70.7) | 105 (73.4) | 1401 (69.7) |  |  | 6294 (71.0) |  |
| Night | 169 (34.6) | 373 (25.9) | 67 (23.5) | 10 (24.4) |  |  | 1274 (29.3) | 38 (26.6) | 610 (30.3) |  |  | 2567 (29.0) |  |
| **Month** |  |  |  |  | <0.0001 |  |  |  |  | 0.0006 |  |  | <0.0001 |
| January | 60 (10.4) | 141 (7.4) | 32 (7.5) | 4 (8) |  |  | 494 (9.7) | 27 (16.3) | 170 (7) |  |  | 928 (8.7) |  |
| February | 56 (9.7) | 115 (6) | 22 (5.1) | 3 (6) |  |  | 385 (7.5) | 10 (6) | 172 (7.1) |  |  | 763 (7.2) |  |
| March | 49 (8.5) | 122 (6.4) | 27 (6.3) | 4 (8) |  |  | 421 (8.2) | 10 (6) | 189 (7.8) |  |  | 822 (7.7) |  |
| April | 32 (5.6) | 128 (6.7) | 29 (6.8) | 3 (6) |  |  | 359 (7) | 6 (3.6) | 169 (7) |  |  | 726 (6.8) |  |
| May | 30 (5.2) | 149 (7.8) | 34 (7.9) | 3 (6) |  |  | 367 (7.2) | 12 (7.2) | 168 (7) |  |  | 763 (7.2) |  |
| June | 52 (9) | 180 (9.4) | 45 (10.5) | 5 (10) |  |  | 479 (9.4) | 15 (9) | 239 (9.9) |  |  | 1015 (9.5) |  |
| July | 38 (6.6) | 193 (10.1) | 50 (11.7) | 9 (18) |  |  | 419 (8.2) | 21 (12.6) | 223 (9.2) |  |  | 953 (8.9) |  |
| August | 27 (4.7) | 131 (6.9) | 23 (5.4) | 3 (6) |  |  | 254 (5) | 10 (6) | 141 (5.8) |  |  | 589 (5.5) |  |
| September | 41 (7.1) | 246 (12.9) | 46 (10.7) | 7 (14) |  |  | 436 (8.5) | 12 (7.2) | 255 (10.6) |  |  | 1043 (9.8) |  |
| October | 64 (11.1) | 199 (10.4) | 37 (8.6) | 4 (8) |  |  | 518 (10.1) | 12 (7.2) | 258 (10.7) |  |  | 1092 (10.3) |  |

|  | Pedestrian<br>(n=575) | Bicycle rider<br>(n=1 908) | Scooter rider<br>(n=429) | Other PMD rider<br>(n=50) |  |  | Car driver<br>(n=5 111) | Truck driver<br>(n=166) | Motorized two-wheelers rider<br>(n=2 414) |  |  | All (n=10 653) |  |
| --- | --- | --- | --- | --- | --- | --- | --- | --- | --- | --- | --- | --- | --- |
|  | n (%) | n (%) | n (%) | n (%) | p-value<sup>1</sup> |  | n (%) | n (%) | n (%) | p-value<sup>2</sup> |  |  | p-value<sup>3</sup><br>(All) |
| November | 72 (12.5) | 179 (9.4) | 41 (9.6) | 5 (10) |  |  | 536 (10.5) | 14 (8.4) | 218 (9) |  |  | 1065 (10.0) |  |
| December | 54 (9.4) | 125 (6.6) | 43 (10) | 0 (0) |  |  | 443 (8.7) | 17 (10.2) | 212 (8.8) |  |  | 894 (8.4) |  |
| **Season** |  |  |  |  | 0.0003 |  |  |  |  | 0.002 |  |  | <0.0001 |
| Spring | 111 (19.3) | 399 (20.9) | 90 (21) | 10 (20) |  |  | 1147 (22.4) | 28 (16.8) | 526 (21.8) |  |  | 2311 (21.7) |  |
| Summer | 117 (20.3) | 504 (26.4) | 118 (27.6) | 17 (34) |  |  | 1152 (22.6) | 46 (27.6) | 603 (24.9) |  |  | 2557 (24.0) |  |
| Autunm | 177 (30.7) | 624 (32.7) | 124 (28.9) | 16 (32) |  |  | 1490 (29.1) | 38 (22.8) | 731 (30.3) |  |  | 3200 (30.0) |  |
| Winter | 170 (29.5) | 381 (20) | 97 (22.6) | 7 (14) |  |  | 1322 (25.9) | 54 (32.5) | 554 (22.9) |  |  | 2585 (24.3) |  |
| **Urban area** |  |  |  |  | <0.0001 |  |  |  |  | <0.0001 |  |  | <0.0001 |
| Lyon | 205 (35.6) | 1257 (65.9) | 325 (75.8) | 26 (52.0) |  |  | 730 (14.3) | 15 (9.0) | 700 (29.0) |  |  | 3258 (30.6) |  |
| Grand Lyon | 220 (38.3) | 498 (26.1) | 77 (17.9) | 18 (36.0) |  |  | 2068 (40.5) | 72 (43.4) | 886 (36.7) |  |  | 3839 (36.0) |  |
| Hors Grand Lyon | 66 (11.5) | 88 (4.6) | 10 (2.3) | 2 (4.0) |  |  | 1375 (26.9) | 53 (31.9) | 388 (16.1) |  |  | 1982 (18.6) |  |
| Rhône SAP | 84 (14.6) | 65 (3.4) | 17 (4.0) | 4 (4.0) |  |  | 938 (18.3) | 26 (15.7) | 440 (18.2) |  |  | 1574 (14.8) |  |

1 Comparison of pedestrian, bicycle rider, scooter rider and other PMD rider
2 Comparison of car driver, truck driver, and Motorized two-wheelers rider
3 Comparison of the 7 categories of injured users

Most PMD riders were under 35 years of age (60% for other PMDs and 52.7% for scooters). Overall, the population was young, with 54.1% aged < 35 years, and 76.1% aged < 45 years. The average age differed significantly according to the user category.

Most work-related scooter crashes occurred during commuting (95.6%). The reason for the journey was related to the mode of travel; 78.9% of the truck driver victims were on duty and 20.7% of pedestrians were involved in the crash while on duty. Overall, 87.2% of the victims were commuters.

Nearly two-thirds of scooter riders and half of other PMD riders had no opponents. Overall, only 26.5% of the victims faced crashes without opponents. By definition, all the pedestrians had opponents.

Of the victims, 90% were involved in crashes during the week, with no difference based on the user category. Less than 25% of scooter and other PMD riders were involved in nighttime crashes. The crashes occurred at night for 29% of the victims, with a higher proportion of pedestrians (34.6%).

The majority of scooter, other PMD, and bicycle riders, and pedestrians were involved in crashes in street-type traffic lanes in the city. More than 45% of car and truck drivers were involved in crashes outside urban areas, whereas approximately 90% of crashes involving scooters, other PMDs, and bicycles occurred in urban areas (Lyon and its metropolitan area).

The use of personal protective equipment according to the user category was the highest for car drivers (97% wore a seatbelt), truck drivers (82% wore a seatbelt), and motorized two-wheeler riders (95% wore a helmet). Only one-third of injured bicycle riders wore helmets, and less than one-fifth of injured scooter and other PMD riders wore helmets.

### 3.2. Description of the injuries

Half of the riders of other PMDs had a single injury, whereas scooter riders, pedestrians, and motorized two-wheeler riders more frequently had multiple injuries (Table 2). Of the victims, 41.9% had a single injury, 33.2% had two injuries, 17.8% had three injuries, and 7.1% had more than four injuries. The average number of injuries differed significantly according to user category (p<0.0001).

**Table 2:** Injury characteristics of victims injured in work-related crash, by user category (source the Rhône Road Trauma Registry, 2015-2020)

|  | Pedestrian<br>(n=575) | Bicycle rider<br>(n=1908) | Scooter rider<br>(n=429) | Other PMD<br>rider<br>(n=50) |  |  | Car driver<br>(n=5111) | Truck driver<br>(n=166) | Motorized<br>two-wheelers<br>rider<br>(n=2414) |  |  | All<br>(n=10<br>653) |  |
| --- | --- | --- | --- | --- | --- | --- | --- | --- | --- | --- | --- | --- | --- |
|  | n (%) | n (%) | n (%) | n (%) | p-value<sup>1</sup> |  | n (%) | n (%) | n (%) | p-value<sup>2</sup> |  |  | p-value<sup>3</sup> |
| **Hospitalization** | 80 (13.9) | 156 (8.2) | 38 (8.9) | 6 (12.0) | 0.0005 |  | 216 (4.2) | 29 (17.5) | 301 (12.5) | <0.0001 |  | 826 (7.8) | <0.0001 |
| **Death** | 3 (0.5) | 2 (0.1) | 1 (0.2) | 0 (0) | NA |  | 2 (0.1) | 1 (0.6) | 3 (0.1) | NA |  | 12 (0.1) | NA |
| **Number of injuries** |  |  |  |  | 0.0001 |  |  |  |  | <0.0001 |  |  | <0.0001 |
| 1 | 186 (32.3) | 759 (39.8) | 163 (38.0) | 25 (50.0) |  |  | 2414 (47.2) | 66 (39.8) | 854 (35.4) |  |  | 4467<br>(41.9) |  |
| 2 | 183 (31.8) | 601 (31.5) | 146 (34.0) | 13 (26.0) |  |  | 1685 (33.0) | 54 (32.5) | 853 (35.3) |  |  | 3535<br>(33.2) |  |
| 3 | 129 (22.4) | 410 (21.5) | 76 (17.7) | 8 (16.0) |  |  | 760 (14.9) | 31 (18.7) | 483 (20.0) |  |  | 1897<br>(17.8) |  |
| 4+ | 77 (13.4) | 138 (7.2) | 44 (10.3) | 4 (8.0) |  |  | 252 (4.9) | 15 (9.0) | 224 (9.3) |  |  | 754 (7.1) |  |
| **MAIS** |  |  |  |  | NA |  |  |  |  | NA |  |  | NA |
| 1 Minor | 451 (78.4) | 1433 (75.1) | 315 (73.4) | 37 (74.0) |  |  | 4830 (94.5) | 138 (83.1) | 1790 (74.1) |  |  | 8994<br>(83.5) |  |
| 2 Moderate | 86 (15.0) | 428 (22.4) | 105 (24.5) | 9 (18.0) |  |  | 223 (4.4) | 22 (13.2) | 516 (21.4) |  |  | 1389<br>(12.9) |  |
| 3 Serious | 21 (3.6) | 37 (1.9) | 6 (1.4) | 3 (0.6) |  |  | 44 (0.8) | 4 (2.4) | 86 (3.6) |  |  | 201 (1.9) |  |
| 4 Severe | 13 (2.3) | 8 (0.4) | 2 (0.5) | 0 (0) |  |  | 10 (0.2) | 2 (1.2) | 14 (0.6) |  |  | 49 (0.4) |  |
| 5 Critical | 3 (0.5) | 2 (0.1) | 0 (0) | 1 (2.0) |  |  | 4 (0.1) | 0 (0) | 8 (0.3) |  |  | 18 (0.2) |  |
| 6 Maximum | 1 (0.2) | 0 (0) | 1 (0.2) | 0 (0) |  |  | 0 (0) | 0 (0) | 0 (0) |  |  | 2 (0) |  |
| **MAIS3+** | 38 (6.6) | 47 (2.5) | 9 (2.1) | 4 (8.0) | <0.0001 |  | 58 (1.1) | 6 (3.6) | 108 (4.5) | <0.0001 |  | 270 (2.5) | <0.0001 |
| **Head injury** | 118 (20.5) | 295 (15.5) | 77 (17.9) | 5 (10.0) | 0.02 |  | 821 (16.1) | 44 (26.5) | 198 (8.2) | <0.0001 |  | 1558<br>(14.5) | <0.0001 |
| **Global gravity of head injury**<br>**MAIS3+** | 11 (9.3) | 12 (4.1) | 2 (2.6) | 1 (20.0) |  |  | 15 (1.8) | 1 (2.3) | 8 (4.0) |  |  | 50 (3.2) | <0.0001 |
| **Face injury** | 73 (12.7) | 315 (16.5) | 91 (21.2) | 7 (14.0) | 0.004 |  | 279 (5.5) | 17 (10.2) | 94 (3.9) | 0.0001 |  | 876 (8.1) | <0.0001 |
|  | n (%) | n (%) | n (%) | n (%) | p-value<sup>1</sup> |  | n (%) | n (%) | n (%) | p-value<sup>2</sup> |  |  | p-value<sup>3</sup> |
| Global gravity of face injury<br>MAIS3+ | 3 (4.0) | 1 (0.3) | 0 (0) | 0 (0) |  |  | 0 (0) | 0 (0) | 0 (0) | NA |  | 4 (0.4) |  |
| Neck injury | 34 (5.9) | 53 (2.8) | 18 (4.2) | 0 (0) | 0.002 |  | 791 (15.5) | 23 (23.9) | 77 (3.2) | <0.0001 |  | 996 (9.2) | <0.0001 |
| Global gravity of neck injury<br>MAIS3+ | 1 (3.0) | 0 (0) | 0 (0) | 0 (0) |  |  | 1 (0.1) | 0 (0) | 1 (1.3) |  |  | 3 (0.3) |  |
| Chest injury | 67 (11.6) | 169 (8.9) | 35 (8.2) | 1 (2.0) | 0.04 |  | 740 (14.5) | 26 (15.7) | 270 (11.2) | 0.0003 |  | 1308<br>(12.1) | <0.0001 |
| Global gravity of chest injury<br>MAIS3+ | 16 (23.9) | 14 (8.3) | 5 (14.3) | 0 (0) |  |  | 31 (4.2) | 5 (19.2) | 52 (19.3) |  |  | 123 (9.4) |  |
| Abdomen injury | 33 (5.7) | 47 (2.5) | 13 (3.0) | 0 (0) | 0.0006 |  | 148 (2.9) | 6 (3.6) | 113 (4.7) | 0.0004 |  | 360 (3.3) | <0.0001 |
| Global gravity of Abdomen<br>injury MAIS3+ | 8 (24.2) | 6 (12.8) | 0 (0) | 0 (0) |  |  | 12 (8.1) | 1 (16.7) | 8 (7.1) |  |  | 35 (9.7) |  |
| Spinal injury | 119 (20.7) | 198 (10.4) | 39 (9.1) | 4 (8.0) | <0.0001 |  | 3114 (60.9) | 70 (42.2) | 373 (15.4) | <0.0001 |  | 3917<br>(36.4) | <0.0001 |
| Global gravity of Spinal injury<br>MAIS3+ | 0 (0) | 2 (1.0) | 0 (0) | 0 (0) |  |  | 6 (0.2) | 1 (1.4) | 5 (1.3) |  |  | 14 (0.4) |  |
| Upper limb injury | 230 (40.0) | 1086 (56.9) | 254 (59.2) | 24 (48.0) | <0.0001 |  | 1121 (21.9) | 55 (33.1) | 1161 (48.1) | <0.0001 |  | 3931<br>(36.5) | <0.0001 |
| Global gravity of Upper limb<br>injury MAIS3+ | 1 (0) | 2 (0.2) | 0 (0) | 0 (0) |  |  | 3 (0.3) | 1 (1.8) | 5 (0.4) |  |  | 12 (0.3) |  |
|  | n (%) | n (%) | n (%) | n (%) | p-value<sup>1</sup> |  | n (%) | n (%) | n (%) | p-value<sup>2</sup> |  |  | p-value<sup>3</sup> |
| **Injury of the lower limbs /**<br>**pelvis** | 395 (68.7) | 899 (47.1) | 201 (46.8) | 26 (52.0) | <0.0001 |  | 750 (14.7) | 45 (27.1) | 1653 (68.5) | <0.0001 |  | 3969<br>(36.8) | <0.0001 |
| **Global gravity of Injury of the**<br>**lower limbs / pelvis MAIS3+** | 18 (4.6) | 23 (2.6) | 4 (2.0) | 3 (11.5) |  |  | 17 (2.3) | 1 (2.2) | 49 (3.0) |  |  | 115 (2.9) |  |
| **New Injury Severity Score**<br>**(NISS)** |  |  |  |  | <0.0001 |  |  |  |  | <0.0001 |  |  | <0.0001 |
| NISS < 9 | 523 (91.0) | 1824 (95.6) | 413 (96.3) | 45 (90.0) |  |  | 5024 (98.3) | 153 (92.2) | 2241 (92.8) |  |  | 10343 |  |
| NISS = [9–15] | 21 (3.6) | 62 (3.2) | 10 (2.3) | 4 (8.0) |  |  | 49 (1.0) | 8 (4.8) | 112 (4.6) |  |  | 266 (2.5) |  |
| NISS ≥ 16 | 31 (5.4) | 22 (1.2) | 6 (1.4) | 1 (2.0) |  |  | 38 (0.7) | 5 (3.0) | 61 (2.5) |  |  | 164 (1.5) |  |

1 Comparison of pedestrian, bicycle rider, scooter rider and other PMD rider
2 Comparison of car driver, truck driver, and Motorized two-wheelers rider
3 Comparison of the 7 categories of injured users

Of the victims, 7.8% were hospitalized. This proportion was higher for pedestrians, drivers of trucks, and riders of motorized two-wheeler and other PMDs. One scooter rider died in an crash. Twelve people died from their injuries (including three pedestrians and three motorcyclists).

The most frequently affected body parts were the spine (37.6%), lower limbs (36.4%), upper limbs (36.1%), head (14.8%), thorax (9.3%), neck (9.7%), face (8.1%), and abdomen (3.3%).

Head injuries were significantly (p<0.0001) more common in pedestrians (20.5%), car (16.1%) and truck (26.5%) drivers and scooter (17.9%) and bicycle (15.5%) riders. Facial injuries were significantly (p<0.0001) more common among scooter (21.2%) and bicycle (16.5%) riders, pedestrians (12.7%), and other PMD riders (14.0%). Neck injuries were significantly (p<0.0001) more common among truck (23.9%) and car (15.5%) drivers. Chest injuries were significantly (p<0.0001) more common among truck (15.7%) and car (14.7%) drivers, pedestrians (11.6%), and two-wheeler riders (11.2%). Abdominal injuries were more common among pedestrians (5.7%) and motorized two-wheeler riders (4.7%). Spinal injuries were significantly (p<0.0001) more common among car (60.9%) and truck (42.2%) drivers, and pedestrians (20.7%) than among other users. Upper extremity injuries were significantly (p<0.0001) more common among riders of scooters (59.2%), bicycles (56.9%), motorized two-wheelers (48.1%), and other PMDs (48.0%), and pedestrians (40.0%). Finally, lower limb injuries were significantly (p<0.0001) more common among riders of motorized two-wheelers (68.5%), other PMDs (52.0%), bicycles (47.1%) and scooters (46.8%).

Only 2.1% of scooter riders were seriously injured (MAIS 3+). Overall, 97.5% of the victims were slightly injured (MAIS <3). The proportion of serious injuries (MAIS 3+) was higher among pedestrians (6.6%), other PMD riders (8%), and motorized two-wheeler riders (4.5%). Differences between global severity and categories of user injured were significant (p<0.0001).

Facial injuries were minor for all crash victims, except for 4% of pedestrians who had injuries of MAIS 3+. Among scooter and other PMD riders, all upper-extremity injuries were of low severity (MAIS <3). However, three riders of other PMD had lower extremity injuries of MAIS ≥3.

### 3.3. Trends in work-related road crashes during 2015–2019

Between 2015 and 2019, an increase in the number of work-related traffic injuries has been observed (28.2%), mainly due to an increase in the number of crashes while commuting (36.3%) and a decrease of the number of crashes on duty (6.9%); 2020, the year of the COVID-19 pandemic, was characterized by a break in this increase (Figure 2).

**Figure 2:**
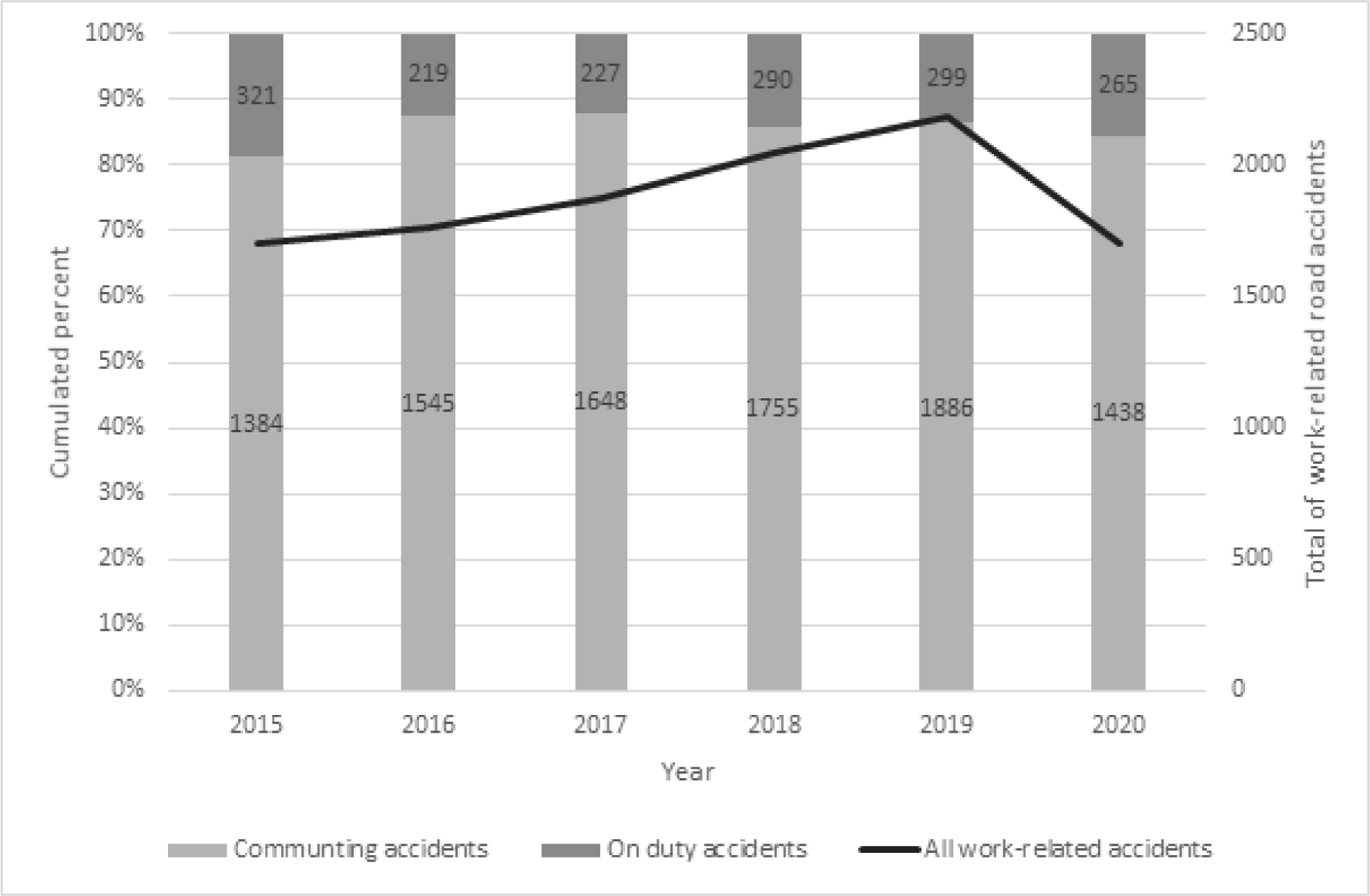
Trends in the number of drivers, riders and pedestrians involved in work-related road crashes, by type of journey between 2015 and 2020 (source the Rhône Road Trauma Registry)

Between 2015 and 2019, the number of scooter riders increased by 773%, that of other PMD by 200%, that of bicycles by 92.1%, that of pedestrian by 33.3% and that of car by 18.3% (Figure 3). In contrast, the number of motorized two-wheeler riders decreased by 5.9%, and that of truck drivers was stable.

**Figure 3:**
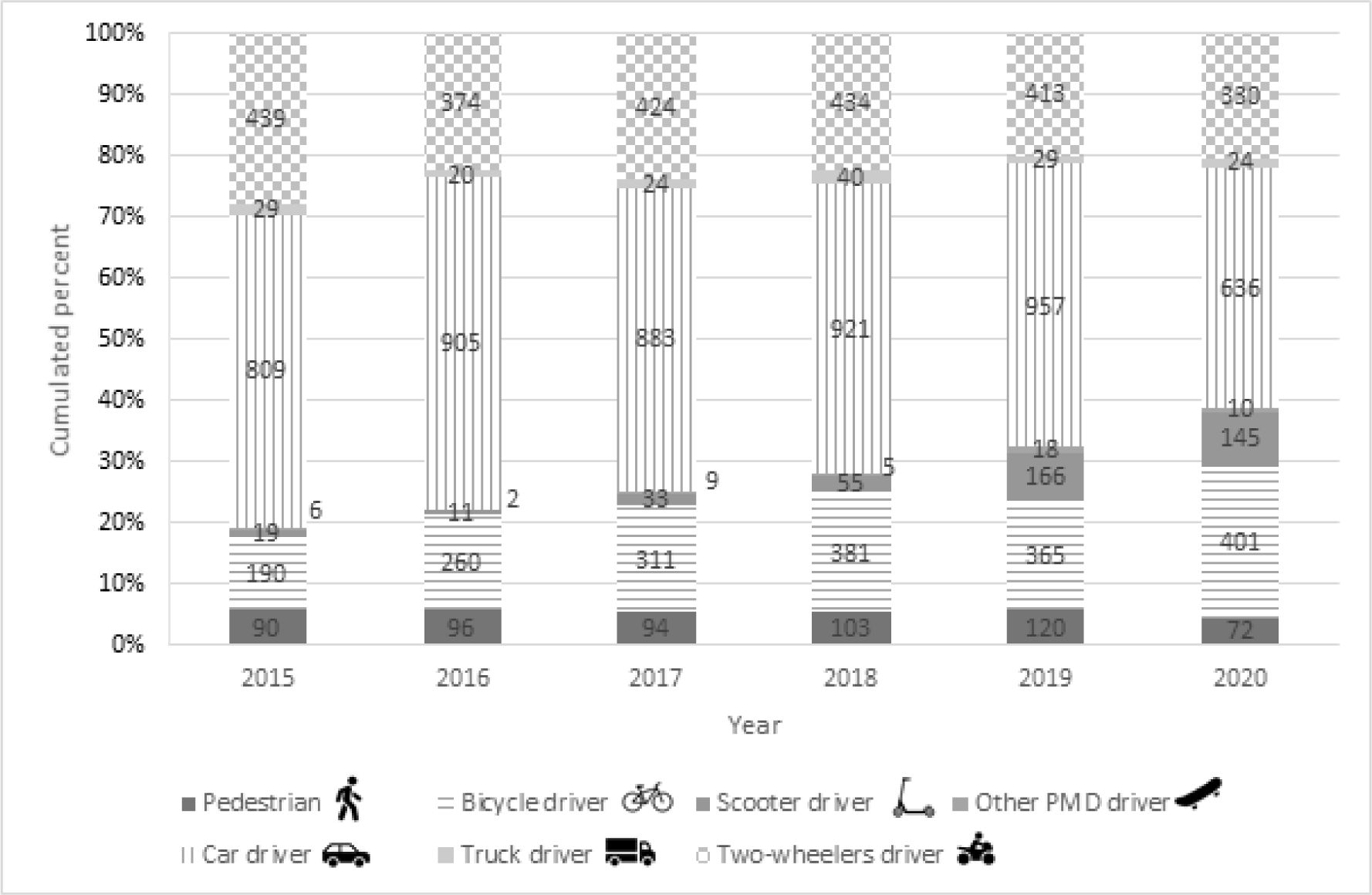
Frequencies (and numbers) of different user categories of drivers, riders and pedestrians injured in work-related crashes, by year (source the Rhône Road Trauma Registry, 2015-2020)

## 4. Discussion

### 4.1. Main findings

This original study on work-related road crashes allowed us to characterize the increase in work-related road crashes associated with new modes of travel, particularly scooters. This new mode of travel has been constantly increasing for approximately 10 years and even more since the emergence of shared electric scooters. Companies in Lyon have offered this service since 2018. Its use accelerated following the COVID-19 pandemic. The results of this study showed that for work-related trips, the proportion scooter riders increased by 773% between 2015 and 2019, and car crashes users increased only by 18.3% in 2020. Bagou et al. [30] found a seven-times increase in scooter crash victims between 2018 and 2019 in the Rhône Department. In contrast, the proportion of car drivers decreased in 2020 due to COVID-19 pandemic-related restrictions [35].

Most work-related road crashes were commuting crashes. Among people working in a fixed and regular place outside the home (i.e., approximately 79% of employed people), approximately 29% make two or more home-to-work trips [2]. This proportion is inversely correlated with commuting journey time. Approximately 25% of employees are exposed to driving as part of their work activities [4,5].

However, only a small proportion of scooter riders (4.4%) were involved in a road crash on duty, while 95.6% of scooter riders had a road crash on commuting. Either their use is reserved for home-work trips or people using this mode of transport have less risky behavior when it comes to a trip within the framework of their professional activity.

### 4.2. Characterization of users of nmPMD/mPMD who had road crash

A prospective study in Spain also characterized electric scooter crashes [36]: women represented 40% of the injured, and approximately 13% of the injured were under 18 years of age. In Austria, two-thirds of the injuries involved men, and 60% of the crashes occurred at night [37]. In Denmark, the majority of injured electric scooter riders were 18-25 years old. The proportion of pedestrians injured by an electric scooter was 17% in crashes involving scooters [38]. A recent German study observed a two-thirds were men, and nighttime crashes [39]. In another study, Graef et al. [40] observed that among patients admitted to a level 1 trauma center with a mean age of 30 years, 44% were female. In Italy, patients involved in electric scooter crashes were predominantly male (79%) with an average age of 30 years [41]. In an annual incidence study of patients with injuries related to the use of electric scooters in the USA, 60% of the victims were men and 92% were riders [42]. In Germany, following the implementation of the first registry to examine injury patterns and collect epidemiological data on people injured while riding electric scooters, Heuer et al. [43] described injuries in 90 patients (65% male; mean age, 35 years).

An early review of the literature on electric scooters provided insights into the circumstances and consequences of electric scooter crashes [44]. The majority of these crashes occur without an opponent and as a result of a fall, collision with an object, excessive speed, or unfavorable road conditions. Nikiforiadis et al. [9] found that electric scooter traffic on sidewalks affected pedestrian experiences the most, ahead of uncontrolled parking on sidewalks. The speed of scooters leads to feelings of insecurity among pedestrians. However, the results of this study showed that scooter opponents were rarely observed among injured pedestrian (12/575). According to a study by Shichman et al. [29], which compared crashes involving electric scooters before and after the introduction of electric-sharing scooters, 80% of the injuries were due to falls. Sixty percent of the scooter riders in our study did not have an opponent and thus, were likely to have fallen in the crash; this proportion was 80% for riders of other PMD.

According to a German study, PMD riders perform secondary tasks while riding [45]. More than 13% of bicycle and electric scooter riders in Germany performed secondary tasks, such as wearing headphones or earphones and chatting with another user. However, the use of cell phones was rare. Riders with a delivery task using a PMD as part of their activities were at higher risk of performing secondary tasks in addition to driving [45]. This observational study found that delivery riders were most likely to wear headphones or earphones; however, they did not show significantly more traffic violations, inappropriate use of roadway infrastructure, or at-fault crashes.

### 4.3. Injuries related to PMD

Most crashes were of low or moderate severity (97.5% MAIS < 3). In the context of work-related road crashes, the frequency of injuries among scooter riders and other PMDs was similar according to the different injury areas and severity. Work-related crashes are generally less severe than private crashes [46].

The upper limbs, lower limbs, face and head were the main areas injured by scooter riders. This finding is consistent with those of other studies. In an Italian study, nearly 47% of the injuries were fractures, with the majority being radial fractures and 25% requiring surgical intervention [36]. In a South Korean study [47], the main anatomical region affected in electric scooter crash victims was the face, followed by the head and upper extremities. In an annual incidence study on patients with injuries related to electric scooter use in the USA [42], the most common injuries were fractures and head injuries. In Germany, among 90 people injured while riding electric scooters, 32 fractures and seven ligament injuries were reported, and head injuries were found in 38 patients [43]. Moftakhar et al. [37] studied the severity of injuries caused by electric scooter crashes in Austria. These crashes occurred at high kinetic energies and caused head and upper extremity injuries. In Germany, following the introduction of shared electric scooters in 2019, the topic of safety emerged as a public debate. A recent study [39] reported that electric scooter crashes were the source of serious injuries, with 28% of crash victims requiring surgical management. In another study, Graef et al. [40] observed that more than two-thirds of patients had extremity injuries, and half had facial injuries. The chest region exhibited the highest AIS scores. Overall, 70% of the injuries were minor. Finally, in Italy, among patients involved in electric scooter crashes [41], 60% had a head injury, 30% were transported to emergency departments for life-threatening injuries, and 15% were placed in intensive care units.

The fact that not all scooter riders used electric scooters in our study minimized the severity of the crashes. Shichman et al. [29] studied crashes involving electric scooters before and after the introduction of shared electric scooters and showed a significant increase in electric scooter-related injuries during emergency department visits after the introduction of sharing services (six times more admissions).

Compared to non-electric scooter riders, electric scooter riders suffered more bruises and lacerations on the face, required sutures, and were more often under the influence of alcohol or drugs [38].

### 4.4. Protective equipment for PMD users

Although seatbelt use among car and truck drivers and helmet use among motorized two-wheeler riders are very high in the context of work travel, the results indicate that helmet use among bicycles, scooters, and other PMD riders is low. This has also been found in other studies in the general population. Helmet use among electric scooter users was anecdotal in a German study [39], only one out of 43 patients wore a helmet in another German study [40], and 4% of victims wore a helmet in a study in Dallas, USA [48].

Helmet use for electric scooter users significantly reduces the risk of serious head and facial injuries [49], maxillofacial fractures, and soft tissue injuries [50]. Similar to the recommendations for helmet use for bicycle users 10 years ago, the scientific literature confirms and recommends helmet use for electric scooter users and, in a larger set, for users of PMDs [51,52]. The non-use of helmets by electric scooter users and the high prevalence of head injuries found in numerous studies [44,53] suggest that these types of injuries may be preventable. Studies demonstrating the effectiveness of helmet use among cyclists can confirm this [54]. As highlighted in this literature review, several cities (Brisbane, Copenhagen, Dallas, Los Angeles, Malaga Paris, Stockolm, and Vienna) and countries (France and Italy) [55,56] have legislated this area [6]. However, results from an Australian study showed that helmet use among commercial shared electric scooter riders was only 60% whereas 95% of private electric scooter riders wore a helmet [57]. It is necessary to ensure that helmets remain available for the commercial shared e--scooters riders or encourage these users to bring their own helmet. Finally, helmet is the least restrictive and most practical form of safety protection.

In order to reduce the risk of injury to upper and lower limbs, several other protective equipments could be used by scooter and other PMDs riders like gloves, jackets with protection or pants. These protective equipments increase the safety of motorized two-wheeler riders. However, it seems difficult to ask scooter riders to wear protective equipment as if they were motorized two-wheelers if wearing a helmet doesn’t seem to be a priority for them already.

### 4.5. Regulations for PMD users

In France, a user of rollerblades, skateboards, or scooters (without a motor) is considered a pedestrian who must ride on the sidewalk. Electrical PMDs (scooters, hoverboards, gyropods, monowheels) must be used on the bicycle path, if available. Bicycles must be on a roadway or bike path, if there is one.

The Mobility Law 2019 [58] established that electric scooters are prohibited on sidewalks (unless authorized by the mayor and unless the engine is turned off). In built-up areas, traffic is only authorized on bicycle paths or, failing that, on roads with a maximum speed limit of 50 km/h. Outside built-up areas, traffic is authorized only on bicycle paths or greenways (except for exemptions). Electric scooters are prohibited for children under 12 years of age. Wearing a helmet is mandatory for minors over 12 years of age and strongly recommended for adults. Civil insurance is mandatory. Riding an electric scooter at speeds exceeding 25 km/h is forbidden. Similar to other means of transportation, the use of headphones is prohibited. Front and rear lights are mandatory, as is wearing a retroreflective vest at night or during low visibility. A horn is mandatory. Finally, passenger transport is prohibited. The regulations mandate fines: 135 euros for driving on sidewalks, 35 euros for failure to comply with traffic laws, and 1,500 euros for exceeding the authorized speed limit [59].

The various regulations on PMDs established by European countries are recent and heterogeneous [60]. Regulations regarding helmet use, specific categories of electric scooters, authorized bike lanes, authorized pedestrian lanes, and age restrictions vary by country. However, the maximum speed limit is a common rule except in Hungary. France and Germany require liability insurance. In the USA, regulations differ by state [56,61]. Overall, many countries are yet to establish the legal requirements of this mode of travel in detail, and more specific legislation is required. In January 2017, the Singaporean government passed the Active Mobility Act (AMA) to regulate the use of PDMs. However, an increase in injuries related to these new travel modes has been observed [62].

Road unsafety related to electric bikes/motorcycles is becoming increasingly problematic owing to their growing popularity compared with other two-wheeled vehicles [63].

The majority of studies on the subject have confirmed that men and users between the ages of 18 and 34 years are most concerned with the use of and crashes involving private or shared PMDs.

In France, one of the few studies on this subject compared the demographic characteristics of users sharing electric scooters with those of personal electric scooters [64]. The latter are more likely to be men, older, and have a higher income. Therefore, the use of electric scooters is motivated by saving time, rather than financial reasons. They also seem to have less risky behaviors, particularly with regard to alcohol consumption, than other electric scooter users [7]. A comparison of the frequency of use (occasional versus frequent users) shows that frequent users have riskier behaviors, perhaps because of their habit of use.

### 4.6. Limitations of the study

Difficulties in correctly identifying nmPMDs were noted. The Rhône Road Trauma Registry collection form evolved in November 2019 to better quantify electric vehicle-related crashes. A “No/Yes” checkbox was introduced for the retrieval of mPMD with certainty, but it does not distinguish nmPMD from missing data.

The analyzed period ended at the start of the COVID-19 pandemic. The year 2020 differed from previous years in that car crashes decreased and scooter crashes increased. It will be relevant to conduct another study spanning a few more years to better measure the impact of nmPMD and mPMD on work-related traffic crashes.

A limitation concerns the geographic area of the study which is the Rhône department of France. Even if data from the Road Trauma Registry are usually used to perform official statistics of road crashes in France [65] because French Police crash data, are incomplete and biased [66], we cannot fully assert that a bias does not exist for work-related road crash in this data.

## 5. Conclusion

- This is the first study describing work-related traffic crashes that have occurred since the emergence of PMDs.
- The results observed for users of scooters and other PMDs in this study were generally consistent with those found in the scientific literature. However, in most available studies, the type of travel was not restricted to work-related journeys: a young and predominantly male population, crashes mostly without opponents.
- Most scooter and other PMD riders had injuries to their upper limbs (59.2%), lower limbs (46.8%), face (21.2%) or head (17.9%).
- Despite limited data, the results suggest that crashes involving scooters or other PMDs are of low severity. Many head injuries could be prevented with more widespread helmet use.
- The use of protective equipment like helmet is essential and not very burdensome and should be mandatory for PMDs and bicycle riders.
- Finally, companies can take preventive actions so that employees using these modes of transport for work-related journeys are better informed about the risks.

## Data Availability

Data may be obtained from a third party and are not publicly available.

## Acknowledgments

We would like to thank all those involved in data collection and computerization, through the Association pour le registre des victimes d’accidents de la circulation du Rhône (Arvac, chaired by Étienne Javouhey) and the Université Gustave-Eiffel-TS2-Umrestte (under the direction of Barbara Charbotel).

## Data Statement

Data may be obtained from a third party and are not publicly available.

## Research ethics

The Rhône Road Trauma Registry has been qualified by the Registry Evaluation Committee (CER) and it has been approved by the National Commission on Informatics and Liberty (CNIL).

